# COVID-19 epidemics monitored through the logarithmic growth rate and SIR model

**DOI:** 10.1101/2021.09.13.21263483

**Authors:** Tomokazu Konishi

## Abstract

**Background:** The SIR model is often used to analyse and forecast the expansion of an epidemic. In this model, the number of patients exponentially increases and decreases, resulting in two phases. Therefore, in these phases, the logarithm of infectious patients changes at a constant rate, the logarithmic growth rate *K*. However, in the case of the coronavirus disease 2019 (COVID19) epidemic, *K* never remains constant but increases and decreases linearly; therefore, the SIR model does not fit that seen in reality. We would like to clarify the cause of this phenomenon and predict the occurrence of COVID19 epidemics.

**Methods:** We simulated a situation in which smaller epidemics were repeated with short time intervals. The results were compared with the epidemic data from 279 countries and regions.

**Results:** In the simulations, the *K* values increased and decreased linearly, similar to the real data. Because the previous peak covered the initial increase in the epidemic, *K* did not increase as much as expected; rather, the difference in the basic reproduction number *R*_0_ appeared in the slope of increasing *K*. Additionally, the mean infectious time *τ* appeared in the negative peaks of *K*. By using the *R*_0_ and *τ* estimated from the changes in *K*, changes in the number of patients could be approximated using the SIR model. This supports the appropriateness of the model for evaluating COVID-19 epidemics. By using the model, the distributions of the parameters were identified. On average, an epidemic started every eleven days in a country. The worldwide mean *R*_0_ was 2.9; however, this value showed an exponential character and could thus increase explosively. In addition, the average *τ* was 12 days; this is not the native value but represents a shortened period because of the isolation of patients. As *τ* represents the half-life, the infectious time varies among patients; hence, prior testing should be performed before isolation is lifted. The changes in *K* represented the state of epidemics and were several weeks to a month ahead of the changes in the number of confirmed cases. In the actual data, when *K* was positive on consecutive days, the number of patients increased a few weeks later. In addition, if the negative peaks of *K* could not be reduced to as small as 0.1, the number of patients remained high. Thus, the number of *K*-positive days and mean infectious time had a clear correlation with the total number of patients. In such cases, mortality, which was lognormally distributed, with a mean of 1.7%, increased. To control the epidemic, it is important to observe *K* daily, not to allow *K* to remain positive continuously, and to terminate a peak with a series of *K-negative* days. To do this, it was necessary to shorten *τ* by finding and isolating a patient earlier. The effectiveness of the countermeasures is apparent in *τ*. The effect of vaccination, in terms of controlling the epidemic, was limited.

## Introduction

An epidemic can be analysed through various approaches, but it is often described using the susceptible-infectious-recovered (SIR) model, which estimates the changes in the number of susceptible (*S*), infectious (*I*), and recovered (*R*) people (Rahimi et al. 2021). This model explains the kinetics by representing the speed of change in the number of corresponding individuals using simultaneous differential equations. This is the most basic mathematical model used in modern epidemiology and is the basis for a family of models that considers more detailed conditions, such as the SEIR model, which includes the exposed (*E*) patients, considering the latency period (Heng & Althaus 2020). An explanation for the SIR model is provided in the Materials and Methods section.

According to the SIR model, the number of infected people increases exponentially until the fraction of susceptible individuals diminishes and then decreases by half at each constant period. In each exponential increase or decrease phase, the logarithms of *I* change linearly over time. The logarithmic growth rate, *K*, indicates the slope of the linear change. Therefore, it should take a constant value in both the stable phases.

However, as far as coronavirus disease 2019 (COVID-19) cases are concerned, *K* increases and decreases in a linear fashion and never remains constant; therefore, the actual cases do not directly fit this model (Konishi 2021a). This conflict raises questions about the use of SIR and related models to understand and predict the status of the COVID-19 epidemic. This could be because variants of severe acute respiratory syndrome coronavirus 2 (SARS-CoV-2) repeat in short intervals with small epidemics that target only a limited population; if the next peak arrives before the previous peak converges, then the early stages of exponential increase will be masked by the previous one. If so, the biphasic pattern is altered. To test this possibility, in this study, simulations were performed and compared with the actual data of the epidemics using exploratory data analysis (Tukey 1977).

Here, we used the basic SIR model exclusively for the following reasons: First, the latency period of the SIER model may differ among patients, but it is rarely measured because the infectious time is difficult to identify (Guan et al. 2020; Lauer et al. 2020; Li et al. 2020). It is also difficult to estimate the number of infections from the data because the effect is indistinguishable from other parameters that affect the speed. By presuming a common half-life of exposed patients, the SEIR model can also be applied; however, this approach increases the assumptions that are difficult to prove (Ellis & Silk 2014). In addition, the latency period in COVID-19 is probably not long because the infectious phase starts before the symptoms appear.

## Materials and methods

### Simulating the SIR model

Here, the model (Rahimi et al. 2021) was modified slightly to correspond to the number of people rather than the percentage. Infection occurs when an infectious person contacts a susceptible person at a constant expectation of infection, *β*, per day. This reduces the number of

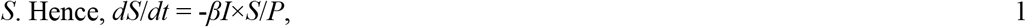

where *P* is the total population. The expectation is *β* = *R*_0_/*τ*, where *R*_0_ is the basic reproduction number, which shows the expected number of each infectious person infected in a suitable condition, *S/P* ≈ 1. *τ* is the mean infectious time; the length was set to five days in the simulations (Alene et al. 2021). The reduced number from *S* represents infectious patients, which will be reduced at a constant rate 1/*τ*,

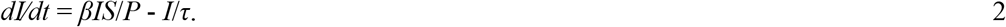

The reduced number represents the recovered individuals as

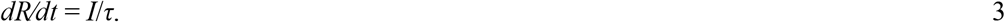

Equations 1–3 represent the model. According to equation 2, when *S/P* ≈ 1 and *S/P* ≈ 0, *dI/dt* becomes a first-order reaction of *I*; hence, *I* increases and decreases exponentially, respectively (IUPAC 2021). Therefore, a peak was formed (Fig. 1A, blue). The R system (R Core Team 2020) was used to simulate the differential equations. The R code used is shown in Figshare (Konishi 2021b).

**Fig. 1.**
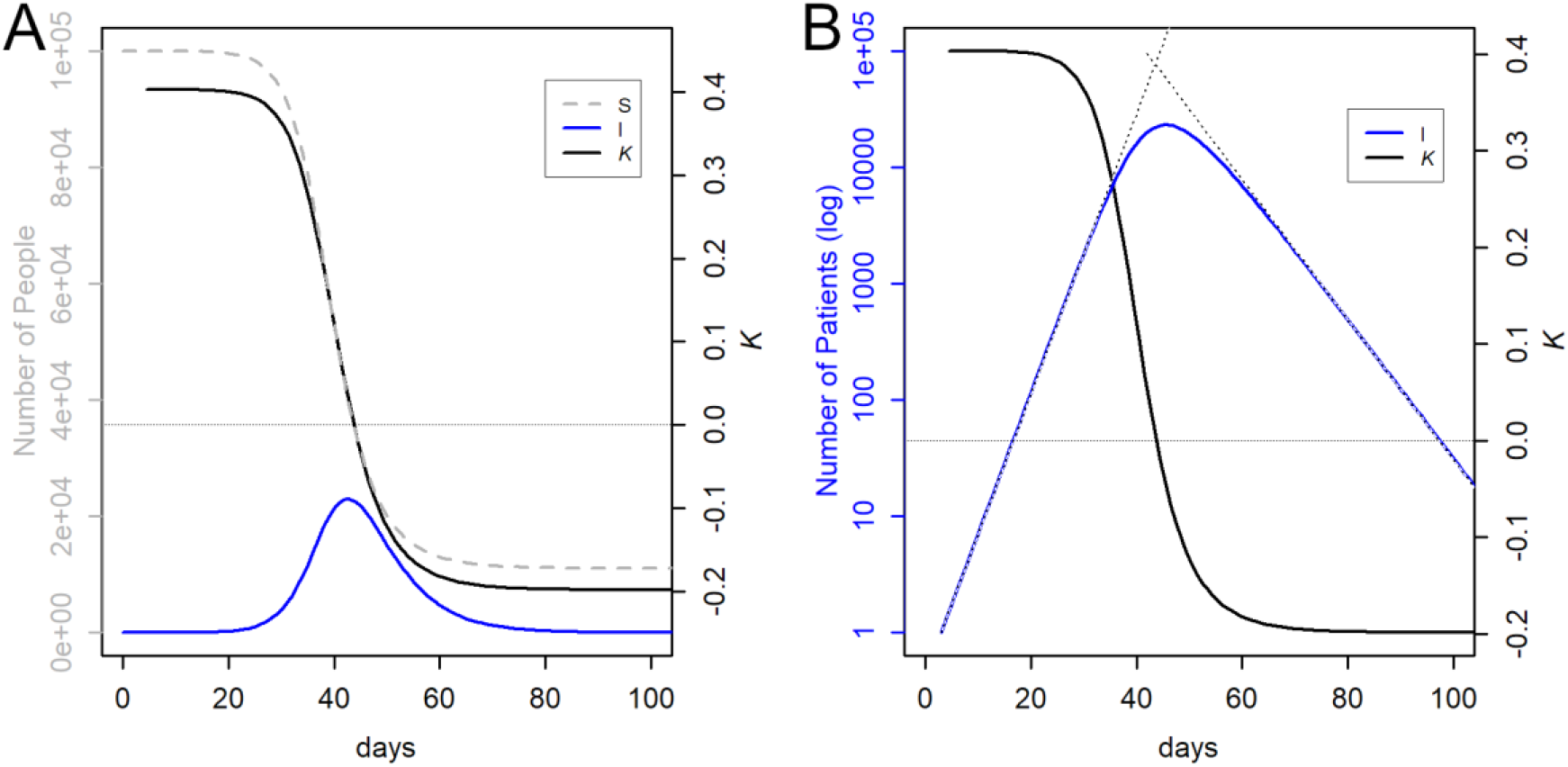
Data simulation using the SIR model. The initial parameters are *S*_0_ = 1E5, I_0_ = 1, *R*_0_ = 4, and *τ* = 5. (**A**) Changes in the number of people. (**B**) *I* in the logarithmic scale. The thin dotted lines are the exponential increase *y* = *R*_0_^*t/τ*^ and decrease *y* = *S*_0_ *×* 2^−(*t*+41)*/τ*^ at *t*th day, respectively. The former means every *τ* day, the number will become *R*_0_ times, and the latter means every *τ* day the number becomes half.

The logarithmic growth rate *K* (Konishi 2021a) is the slope of the logarithm of the exponential change *I* (Fig. 1B, blue). Because the slope is constant, *K* should also be the same (black). Here, it is defined as *K* = *d* (log_2_ *I*)*/dt*. Because 2 was used as the base of the logarithms instead of *e*, 1/|*K*| shows the doubling time (*K* > 0) or half-life (*K* < 0), directly. *R*_0_ is a value that depends on the SIR model and is affected by *τ*, while *K* is a more physically determined parameter that is independent of the model and valid as long as the subject changes exponentially. When *S/P* ≈ 0 (*K* < 0), the number of patients after *t* days will be 2^*-t*/*τ*^ = 2^*Kt*^ times. This results in *τ* = 1/(− *K*). When *S/P* ≈ 1 (*K* > 0), the number of patients after *t* days becomes *R*_0_ ^*t*/*τ*^ = 2^*Kt*^ times. This results in *R*_0_ = 2^*Kτ*^.

Note that when *R*_0_ is low, the exponential infection stops, leaving some *S*_0_ uninfected. From equation 2, we obtain *dI*/*dt* = (*βS*/*P* - 1/*τ*) *I*. 2’

When *βS*/*P* - 1/*τ* > 0, an exponential increase was observed. Because *β = R*_0_/*τ*, this can be transformed to *R*_0_ *> P*/*S*. This results in *S* > *P*/*R*_0_, which shows the limit of the exponential increase, but this does not directly represent the number of people who can escape the infection, as the infection may continue after the exponential increase and vice versa. For example, the simulation showed that 70% and 20% of people may be left uninfected when *R*_0_ is 2 and 3, respectively (Fig. S1).

Because the simulation results in exponentially varying outcomes, the calculations were somewhat unstable, resulting in differences between the input and output parameters. Therefore, we used the calculation results (Fig. 1, black) to estimate *R*_0_ and *τ* values in the simulation; for example, in the case of Fig. 1, *K* took two constant phases at 0.4 and -0.2; hence, *τ* = 1/-(−0.2) = 5 and *R*_0_ = 2^0.4*5^ = 4. The instability was especially apparent in the condition of input *R*_0_ < 2. Under such conditions, the exponential increase was likely to stop, leaving many *S*_0_ uninfected (Fig. S1), and the computational values of *τ* and *R*_0_ tended to increase. This is a kind of artificial error, but unfortunately, it was unavoidable even when using alternative codes, such as the deSolve package (Soetaert et al. 2010). Thus, *R*_0_ << 2 is difficult to reproduce.

The closer the peaks are to each other or the wider they are, the negative peak of *K* is higher (Fig. 2B and 2C). This effect was simulated as follows. For a given *R*_0_, we set various mean infection times and estimated a single peak of *I* using the SIR model. Bimodal peaks were artificially synthesised by superimposing the peaks at intervals of 40 d. As discussed later with real data, this interval is longer than the ordinal condition but is a possible length. The negative peak sandwiched between two identical peaks was measured, and *τ* was estimated as 1/(-*K*) and then compared to the original constant phase. It was confirmed that *R*_0_ did not move significantly during the simulation to change *τ*. At each *R*_0_, simulations were performed until the peaks were too close together, and the valleys were no longer observed.

**Fig. 2.**
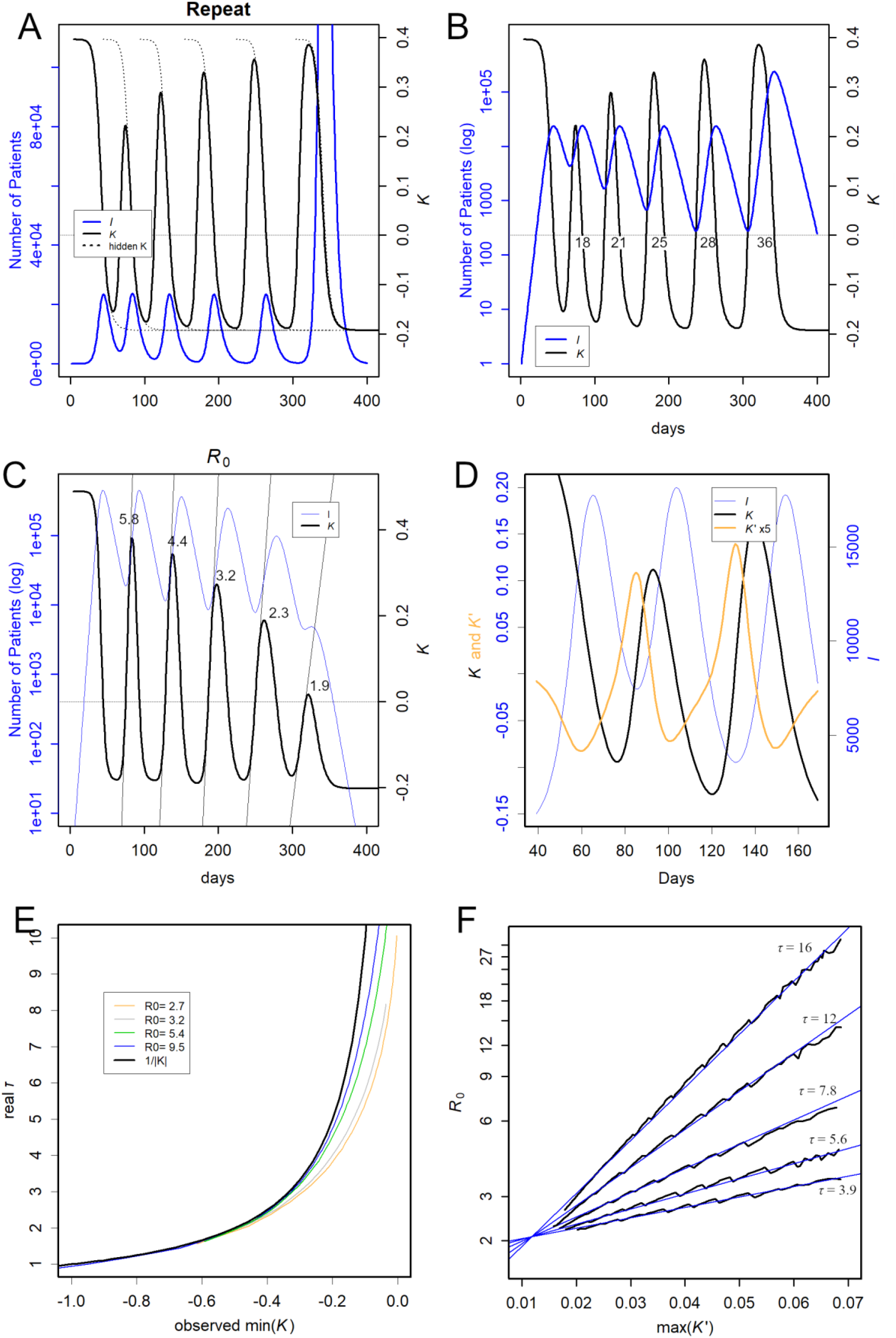
Simulation of repeated epidemics. (**A**) New epidemics started after 40, 90, 150, 220, and 290 days from the first one. Each epidemic started from I_0_ = 1 and *S*_0_ = 1E5, but *S*_0_ = 1E6 was observed only for the last time. (**B**) Semi-log display of *I*. The numbers indicate the days when *K* was positive. (**C**) Infection was initiated every 40 days at the indicated *R*_0_. The thin solid line indicates the slope of *K*. (**D**) Comparison of *K* and *K*’. The peak of *K’* is near the middle of the upward slope of *K*. (**E**) Relationship between the observed negative peak of *K* and the mean infectious time, *τ*, of the used data. 1/|*K*| (black), which is used for the estimation of *τ*, is always larger than that in reality (coloured). (**F**) Relationship between the peak of *K’* and *R*_0_ estimated by simulations at the *τ* presented. A semi-log plot. Blue straight lines present the estimated relationship deduced from *τ* (Fig. 3); these are not the regression lines.

We simulated the effect of *τ* on the estimation of *R*_0_ from the peak of *K*’ ≡ *dK/dt*. A series of *R*_0_ values were set under a certain *τ*, and the peak of *I* was estimated using the SIR model. These peaks were superimposed 20 days after the peak at *τ* = 5 and *R*_0_ = 5. The *K* and *K*’ values were calculated from the synthetized bimodal peaks, and the slope of the rising edge of the latter *K* peak was estimated from the peak of *K*’. This *K*’ value was compared with the *R*_0_ of the latter peak, estimated in the second constant phase.

### Epidemic data

Data on the number of infected people and fatalities were obtained from the Johns Hopkins University repository (Dong et al. 2020) on 01 September 2021. These values for Japan were from the government’s site (Ministry of Health, Labour and Welfare 2021). In the actual data, *I* represented the daily confirmed cases; as they fluctuated, a moving average of 9 days interval was used. *K* was calculated from the difference in the moving average over a 7-day period, to avoid the influence of the day of the week, and represented the moving average of the 9 days interval. The mortality rate was calculated as the number of deaths after 7 days per number of patients on a particular day. The moving average of the 9-day interval was used.

### Finding peaks and estimation of *R*_0_ and *τ*

The peak of *K*’ was found in the following way: the peak of *K*’ occurred when *dK*’/*dt* changed from positive to negative (Fig. S2A). The maximum *K’* during the four days before and after this change was recorded as the peak day and peak height. In practice, *dK*’/*dt* fluctuates; therefore, we used a 9-day moving average to calculate it. The intervals of the peaks were estimated using peak dates. The negative peak of *K* was also detected in the same way using *K*’, and the peak heights were used to estimate *τ*. As shown in Fig. 4B, the regression was effective at -0.04; however, only values less than -0.08 were used to avoid the noise of interference for safety. By using the *K*’ peaks and *τ*, a series of *R*_0_ values in a country was also estimated using the equation presented in the legend of Fig. 3.

**Fig. 3.**
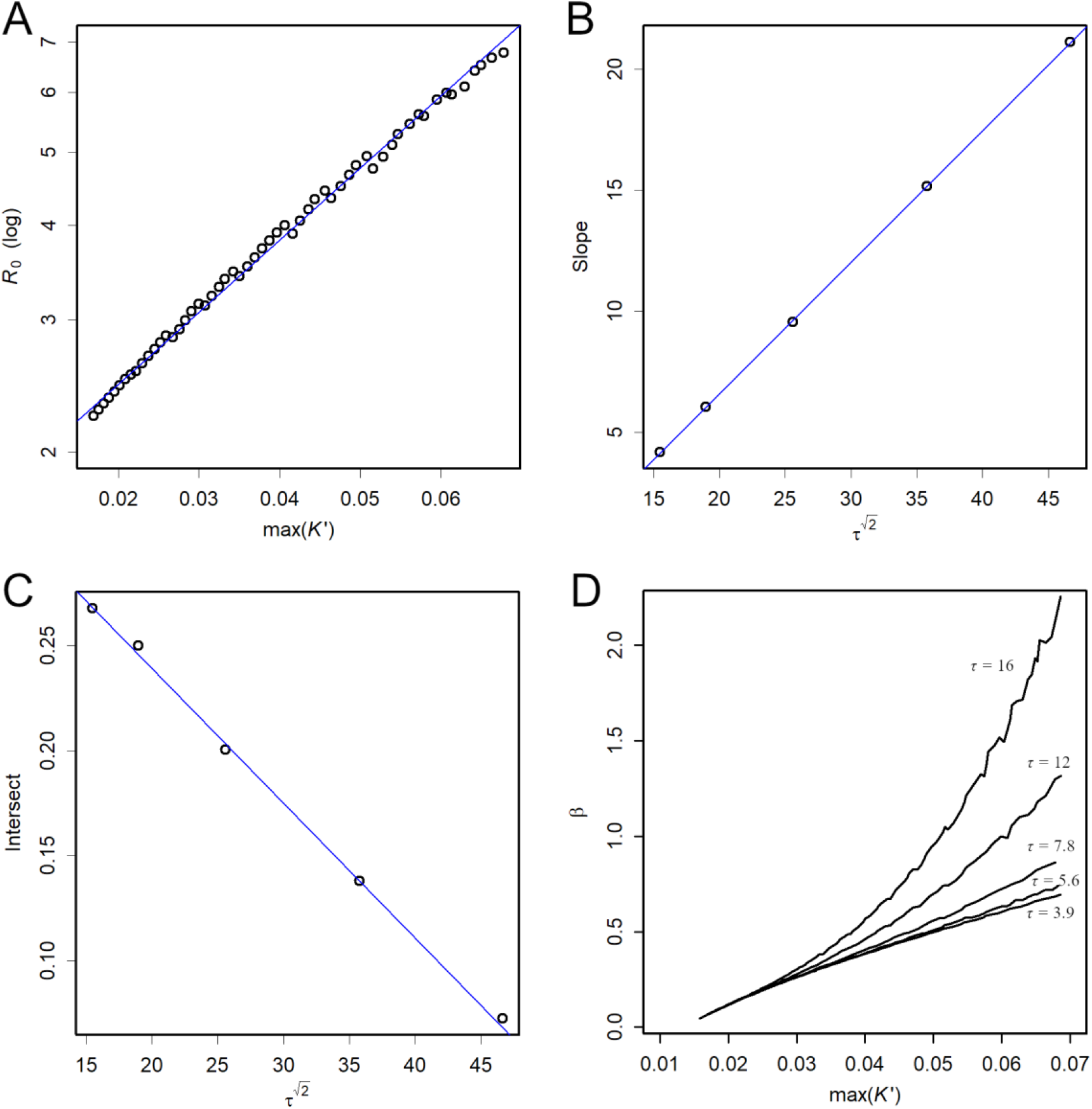
Relationships between the mean infectious time *τ* and other parameters. (**A**) Simulated relationship between the peaks of *K*’ and *R*_0_. Here, the regression line was robustly estimated by the line function of R (Tukey 1977). (**B**) Relationship between *τ* and the slope of the regression line in (A). The slope is *a* = *τ* ^(2^0.5)*0.5436 - 4.2803. (**C**) Relationship between *τ* and the intersect of the regression line in (A). The intersect is *b* = *τ* ^(2^0.5)*(− 0.006406) + 0.367251. These values were used in estimating the relationships in Fig. 2F. (**D**) Simulated relationship between *K*’ peak and *β*. When *τ* is small the relationship is almost linear, while this would likely become exponential when *τ* is larger.

**Fig. 4.**
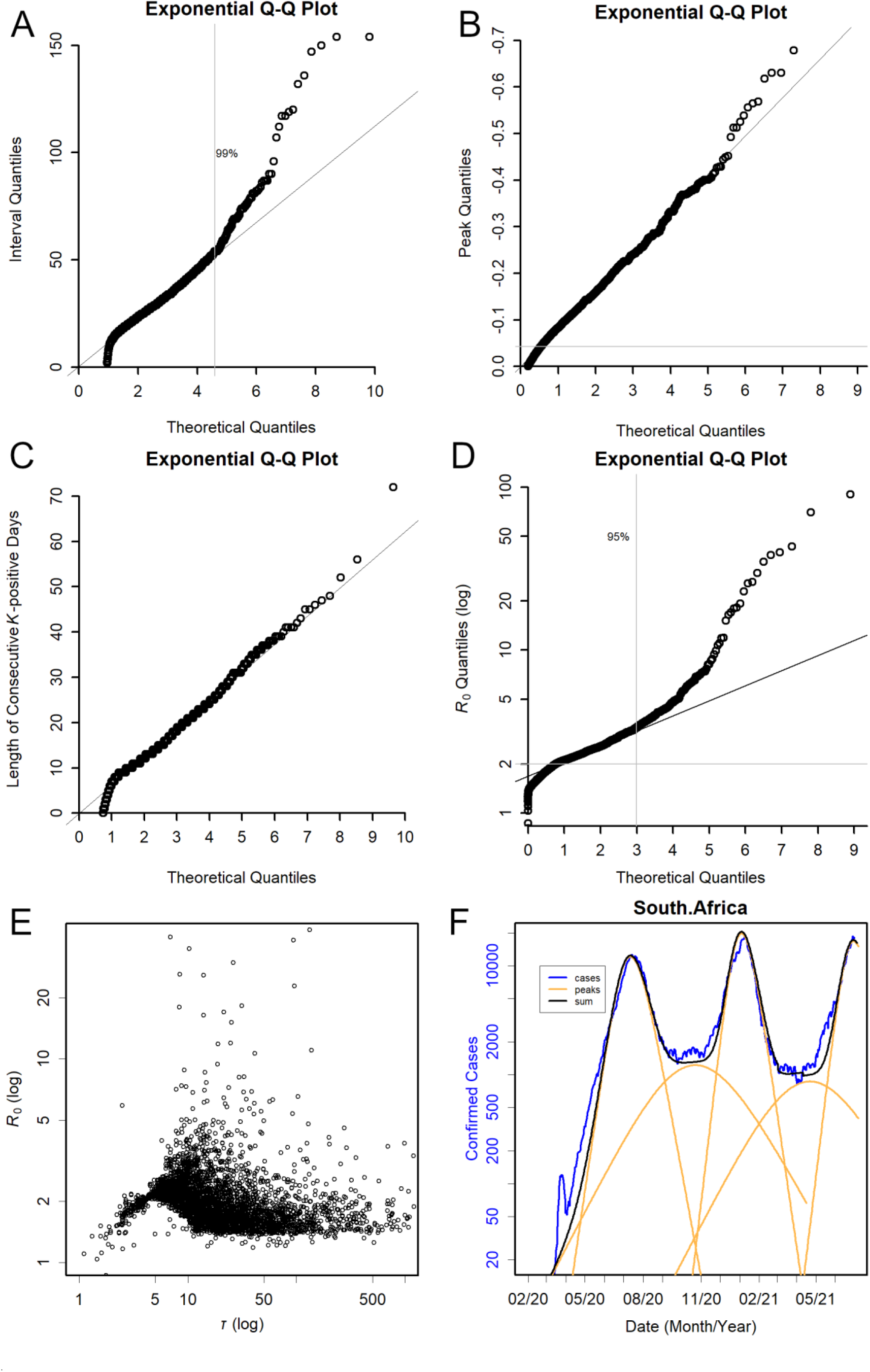
Distributions of peak-related values. (**A**) Correspondence of quantiles of the intervals between the peaks with that of the exponential distribution. If data obey this distribution, a straight line is observed. The slope of the regression line was 11.3; this equals the mean and standard deviation of the distribution. The vertical grey line, which presents the upper limit of coincidence with the theoretical values, shows the percentile indicated. (**B**) Negative peaks of *K*. The horizontal grey line shows the upper limit of linear correlation; note that the y-axis is reversed. (**C**) Consecutive K-positive days. The slope was 6.2 days. (**D**) Estimated values of *R*_0_, semi-log plot. (**E**) Relationship between *τ* and *R*_0_. When *τ* were small, *R*_0_ was always small, and when *R*_0_ was large, *τ* was large. (**F**) Approximation of the confirmed cases by using estimated *R*_0_ and *τ*.

The changes in *I* of a country were approximated by the SIR model using the least number of peaks. The number of confirmed cases in South Africa had three obvious peaks; in addition to these three, two concealed peaks were temporarily placed in between. The *S*_0_ of each peak was estimated from the fragments of data that were roughly dissected vertically. For the three obvious peaks, *R*_0_ and *τ* were estimated from the peaks of *K*’ and negative peaks of *K* (Fig. 4F); for those of the two minor peaks, which were a combination of the smaller peaks, the average values were used. The increase and decrease in each of the peaks were estimated using the SIR model and summed. As *I* of the real data presents the number of confirmed cases in the day, the simulation data for *I* will become larger for *τ* because this is the total number of days for *τ*. Hence, the number d*S*/d*t is* used instead.

### Quantile-quantile (QQ) plot

The quantile-quantile (QQ) plot compares the quantiles of data with those of a particular distribution; this was done to find a suitable model for the data (Tukey 1977). The peak heights, peak intervals, *R*_0,_ and mortality rates were determined. For the theoretical values, the exponential distribution, which is frequently used to describe intervals of randomly occurring events and the normal distribution (Stuart & Ord 2010), was tested. By comparing the quantiles of the real data and the theoretical value, a linear relationship was obtained if the data followed the distribution model. It should be noted that because the mode of the exponential distribution is in the lowest class, the plot at the upper classes becomes thin (Fig. S3A).

There is a limit to the resolution of the short intervals of peaks; two peaks that are too close are counted as one. Therefore, the short intervals were neglected. This is problematic because the mode of the exponential distribution has the smallest values; in fact, it is named because the probability density function decreases exponentially. When there are such missing intervals, the regression line of the QQ plot does not pass through the origin, although the smallest interval should be zero. The distribution of the data was evaluated by compensating for this missing data by adding a set of arbitrary negative values to the data so that the line passed through the origin. The added values do not appear in the plot; therefore, the missing data will create a space in the smallest area. As a property of exponential distribution, such compensation does not change the shape of the distribution on the QQ plot, but only shifts it horizontally in parallel. Therefore, the slope, which represents the mean, was maintained.

## Results

In a simulation of a single epidemic observed alone, *K* inherently showed biphasic constant values (Fig. 1, black). In this case, *K* became constant at 0.4, when *S/P* ≈ 1, and at -0.2, when *S/P* ≈ 0. As a result, *I* increased and decreased exponentially; therefore, the changes became linear when considered in logarithmic form (Fig. 1B). The dotted lines in Panel B show an exponential increase and decrease with the estimated constant rates, respectively.

However, in a real-world scenario, *K* increases and decreases linearly, never becoming constant (Konishi 2021a). In the simulation, such a linear upward and downward trend was observed when the peaks had overlapping tails. Fig. 2A shows the cases where the infection from a new strain started after 40, 90, 150, 220, and 290 days from the first one. Each *S*_0_ was 1E5, but only the last peak was given 10 times the number of people. The constant phase disappeared because the previous peaks masked the increase in the earlier days of *I*, when *I* was still small; the original *K* of the infections that started late are shown by the dotted lines (Fig. 2A).

The movement of *K* precedes the movement of *I* by several weeks (Fig. 2). The exponential increase begins before *K* turns upward, but this turning occurs several weeks before *I* begins to rise visibly. Similarly, we have to wait several weeks after *K* shows a downward trend before *I* actually decreases; when *K* becomes positive, *I* is in the valley bottom between peaks; when *K* becomes negative, *I* is at the peak top.

The peak top of *K* decreased as the peak came closer to the previous peak. The period when *K* is positive (indicated by the numbers in Fig. 2B) also changes; the closer it is to the previous peak, the shorter it becomes. It should be noted that the total number of *I* became ten times higher just by increasing this period from 28 to 36 days.

Compared to the sensitive change in the peak tops because of the overlapping peaks, the negative peak did not change significantly (Fig. 2B), which also appeared when the width of the peak was changed by altering *R*_0_ (Fig. 2C). This may allow the estimation of the mean infectious time from the negative peaks of *K*, as 1/(-*K*). Of course, if the peaks are close together (Fig. 2B) or the widths of the peaks are wide (Fig. 2C), interference will occur, and a negative peak is observed at a higher position than in the original biphasic state. However, if there is a period when the peak interval is sufficiently large to create a window of visibility, negative peaks of *K* can be observed. The effect of this interference between peaks was confirmed by simulations with various *τ* over a set of twin peaks of the same size with a 40-day interval (Fig 2E). As shown below in the real data, an interval of this magnitude can be expected in a few months (Fig. S3B). Estimations using *τ* =1/(-*K*) (Fig 2E, black) are always larger than the actual *τ* (coloured); thus, it is a safe method of estimation. Therefore, the *K* observed at a small level is appropriate for estimating the level of *τ* in the country. Additionally, a heavy interference is visibly apparent; hence, it can be simply eliminated, although the simulation was performed until just before the two peaks overlapped and became one.

The difference in *R*_0_ appears in the slope of *K*. Fig. 2C is the result of simulating the epidemic at the presented *R*_0_, in which the epidemics started at 20-day intervals. The slopes may stably present *R*_0_ compared to the peak tops (Fig. 2B), and we can use the peak tops of *K*’ to define the slope (Fig. 2D). However, since *R*_0_ = *βτ*, the value of *R*_0_ depends on *τ* at that time, even if the variants have similar infectivity. Because *R*_0_ = 2^*Kτ*^, *R*_0_ may intrinsically change its value exponentially. The simulation showed that the value *R*_0_ changed exponentially according to the slopes, and the difference in mean infectious time, *τ*, alters the estimation of *R*_0_ (Fig. 2F).

The relationship between *τ* and other parameters can be predicted empirically, although this relationship was not solved analytically. Fig. 3A shows the relationship between the *K*’ peak and the logarithm of *R*_0_ for *τ* = 9.9, showing that they have a linear relationship over a wide range. Linearity was also observed for different values of *τ* (Fig. 2F), but the slope and intercept varied with *τ*; they showed a linear relationship defined as 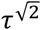 (Figs. 3B and 3C). Therefore, when *τ* is available through the negative peak of *K*, we can translate the peak of *K*’ to *R*_0_ by using only *τ* (the legend in Fig. 3). The lines in Fig. 2F are not regression lines but are relationships estimated from *τ*, showing agreement with the simulation. The length *τ* also affects the relationship between *K*’ and *β* and the expectation of infections per day (Fig. 3D).

Here, we consider actual data by using the Johns Hopkins University repository (Dong et al. 2020) which covers data from 279 countries and regions. First, the intervals between peaks followed an exponential distribution (Fig. 4A), which represented the intervals of randomly occurring events (Stuart & Ord 2010). As the shorter intervals may have been missed, this distribution compensated for the missing data (Materials and Methods). The data in the linear range of the QQ plot were probably generated by a common mechanism. Data outside this range are likely to be affected by some effects, including noise. The top 1% were above the regression line; these longer periods were reported in well-controlled countries, where peaks were rare. The slope of the regression line was 11.3 days; as a characteristic of the exponential distribution, this is the mean and standard deviation. According to this distribution, the 40-day interval corresponded to the 97th percentile (Fig. S3A), with a frequency of approximately once every few months (Fig. S3B).

When the peaks of *K* and *K*’ were observed for all the countries, they were also distributed according to the exponential distribution (Fig. 4B and S2B). Although the peaks of *K’* and the negative peaks of *K* were detected from both sides of the negative and positive peak heights, all the data were used as is. Incidentally, the *K* and *K*’ peaks appeared to be unrelated to the interval length (Fig. S4); hence, the distribution was not determined by the intervals. There were approximately 3600 peaks in *K*’ and *K* between 2020-05-01 and 2021-07-01. Although many peaks may have been missed (Fig. 4A), the distribution was clearly observed, suggesting that the missing peaks occur randomly, possibly because of the short intervals, and are not related to the peak height.

In the negative peaks of *K*, from which the mean infectious time could be estimated as *τ* = 1/(-*K*) (Fig 2E), the slope was -0.082; hence, the mean of *τ* was 12 days (Fig 4B). The 10th percentile of the data was -0.2, representing 5 days, which is close to the value reported in the meta-analysis (Alene et al. 2021). The grey horizontal line represents the upper limit of the linear relationship, which corresponds to *K* = -0.04. Data with larger values may be heavily affected by noise (Fig. 2E).

The number of consecutive *K*-positive days was exponentially distributed (Fig. 4C). It is possible that shorter intervals were missing; hence, this was compensated for. The average was 6 days, but the mode observed was 10 days.

A series of *R*_0_ values in a country was estimated using the peaks of *K*’ and estimated *τ*. Because *R*_0_ is calculated as *R*_0_ = 2^*Kτ*^, the logarithms of *R*_0_ were compared with the theoretical value of the exponential distribution, although the relationship bent slightly downward (Fig. 4D). The top 5% of the data had higher values than the regression line. The ratio was larger than the slope upward of *K*’ (Fig. S2B); therefore, this may include the effect of the extension of *τ* in some countries, in addition to that of super-spreaders and the newest infectious variants. In fact, *τ* affects *R*_0_; when *τ* is small, *R*_0_ remains low, and when *R*_0_ is large, *τ* is always large (Fig. 4E). The high values of *τ* more than 1/-(−0.04) = 25 may be affected by noise (Fig. 2E and 4 B); reduction of the noise by compressing Fig. 4E to the left would show a monotonic increase. When *K* could be reduced to - 0.1 and, hence, *τ* was less than 10 days, *R*_0_ could be maintained at a low level (Fig. 4E), which may be less than the limit level of exponential increase (Fig. 4D and S1A).

Many of the smaller data in Fig. 4D may have been heavily affected by noise in measuring the slopes (Fig. 2F); thus, the plot turned downward. The lower limit of the regression was approximately *R*_0_ = 2; measurements less than this level would be inaccurate because of noise. Since the missing data probably occurred independent of the magnitude of *R*_0_, they were not compensated for. Rather, this distribution should be considered to have a positive minimum value, the background; here, it was 1.7, that is, the intersection of the regression line. This extrapolated value could be the least value that could enable an exponential increase, which would result in detectable peaks of *K*. The slope was 0.1; hence, the mean of the basic reproduction number was *R*_0_ = 1.7+10^0.1^ = 2.9. Owing to the nature of the exponential distribution (Fig. S3A), values smaller than 2 were frequently observed (Fig. 4D). For safety, these values should be recognised as “less than 2 and larger than 1.7”.

Using the estimated *R*_0_ and *τ*, the confirmed cases from South Africa were approximated by using the SIR model using the least number of peaks estimated: three obvious peaks and two in between them (Fig. 4F). The match is particularly close for the three obvious peaks, indicating that the estimates of *R*_0_ and *τ* were reasonably accurate. Here, 21 peaks of *K* were identified; it would be possible to approximate the results with more accuracy by using all these peaks; however, estimations of the location of each peak and the assignment of *S*_0_ would require fine tuning, such as an approach of repeating simulations to find the optimal solution.

Below is a snapshot of the situation in a few countries (Figs. 5–7). The full set of results is shown in Figshare (Konishi 2021b). In almost all cases, *K* always increased or decreased linearly. Note that the scales of *K* and *K*’, presented on the left-hand axis, are common among all the figures, but the number of confirmed cases on the right-hand axis varies significantly.

**Fig. 5.**
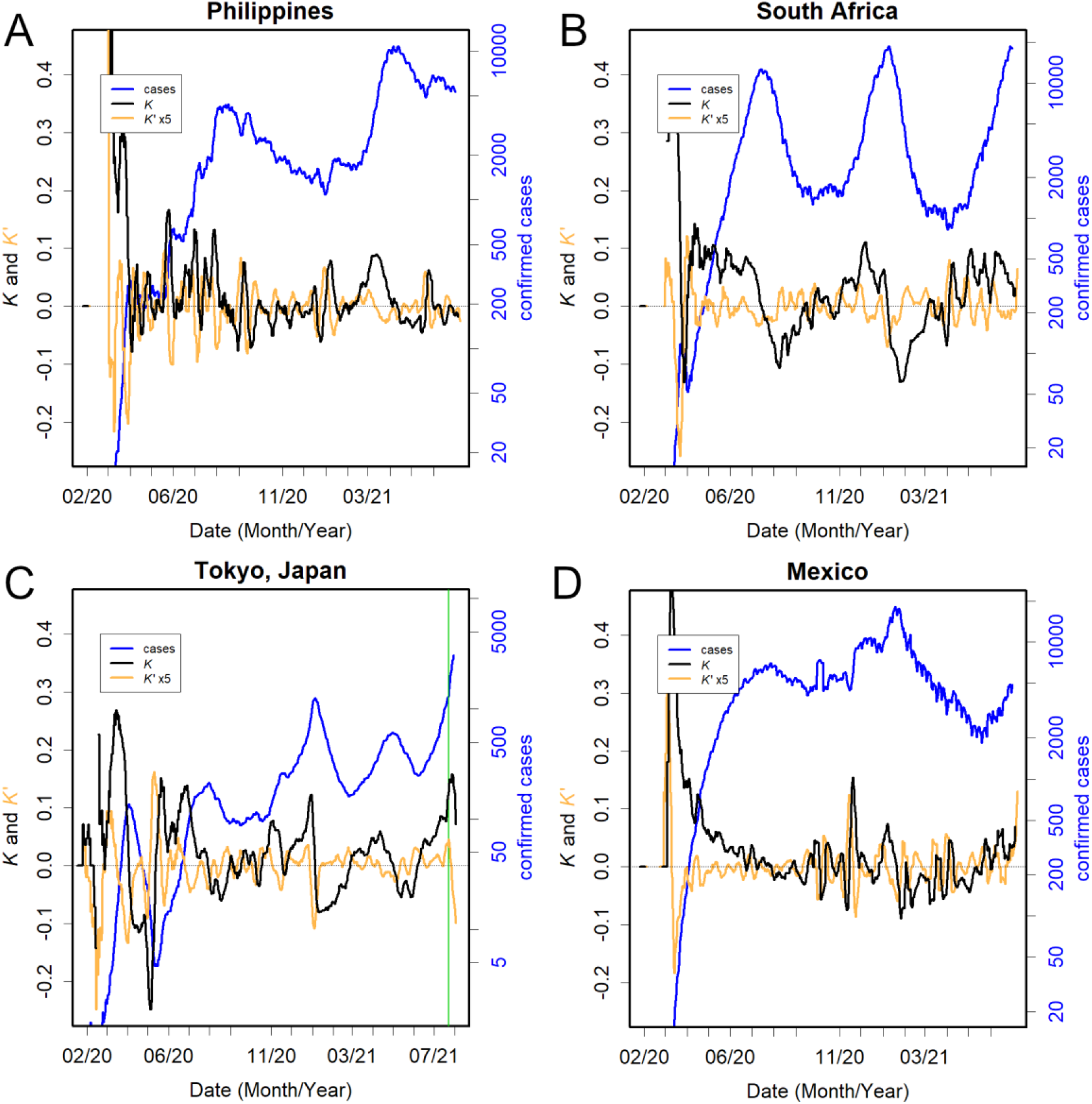
Actual data, a typical example of continuum of positive *K*. (**A**) The Philippines, (**B**) South Africa, (**C**) Tokyo (Japan), and (**D**) Mexico.

When the variant is replaced by a new one, the previous measures lose their effectiveness, people with certain lifestyles become the new targets as *S*_0_, and a new epidemic starts. This phenomenon is evident in Figs. 5 and 6. The Philippines is an example that has two opposite aspects: success and failure of controlling epidemics (Fig. 5A). Until April 2020, *I* remained low because there was no *K-*positive continuum. However, in May, when the variant changed, the *K* positives became consecutive, and a large peak was reached. Thereafter, while the series of *K-positives* was halted, the negative days could not be controlled and *I* continued to remain high, creating a large peak when a new variant came in January 2021. New variants produced peaks in South Africa and Tokyo (Fig. 5B and 5C) (Konishi 2021a; Konishi 2021c). As *I* did not drop completely, a few *K-*positive days in a row led to an explosion of the infection. Several countries showed this trend, and Mexico was an extreme example (Fig. 5D).

**Fig. 6.**
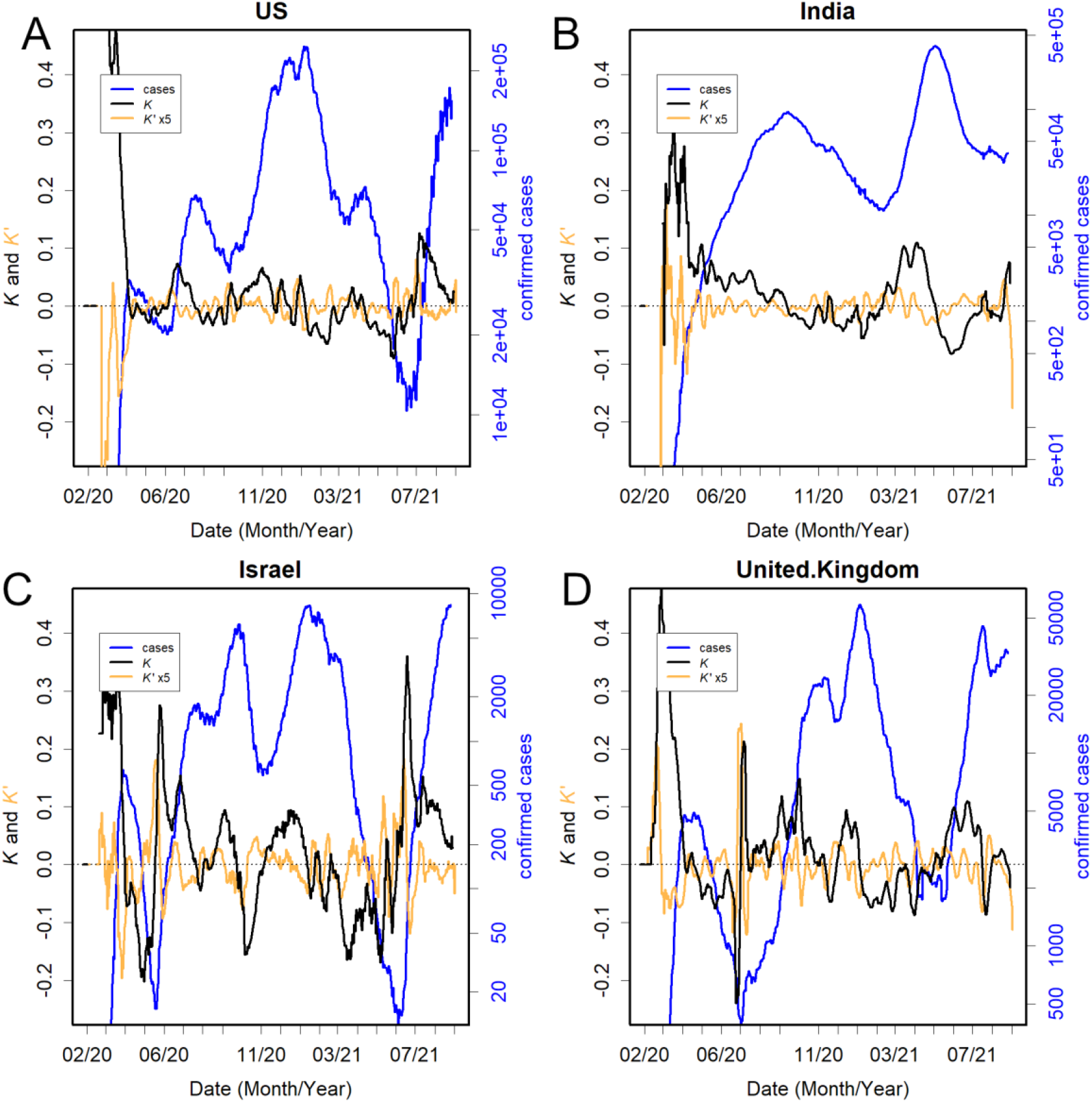
Countries with long-positive *K*. 2. (**A**) USA, (**B**) India, (**C**) Israel, and (**D**) UK. Vaccination coverage (2 doses) in these countries was 47%, 4%, 57%, and 49%, respectively (Our World in Data 2021, P 5 July).

Although the Olympics were held in Tokyo, the mean infectious time in this city, in June 2021, was 16 days. This shows that the measures are not working well and that the delta variant (WHO 2021) is spreading (Fig. 5C). Since the opening of the Olympic Games (green vertical line), there has been an explosion of infections in this city, with an *R*_0_ of 15. This is one of the worst data in the world to date (Fig. 4D).

Other disastrous examples of repetitive *K-*positives were the US and India (Fig 6 AB). *K* increases not only when variants are new, but also when there is something new about the way people live. There were large elections in these countries. Politicians did not ask people to self-restrain; instead, they mobilised defenceless people for election rallies (Menon & Goodman 2021; Norris & Gonzalez 2020). There was also a major religious event in India (Pandey, 2021). These countries allowed *K*-positive days to last for unusually long periods, producing the world’s first and second largest cumulative number of cases. It should also be noted that in both countries, all the series of *R*_0_ detected were less than 2.

Another characteristic of these countries is that their mean infectious time was long. For example, in the US and India, the *K* negative peaks were greater than -0.05 (Fig 6 AB, orange); therefore, the mean infectious times were probably longer than 3 weeks. The other heavily infected countries seem to be the same (Fig. 5 and 6).

Vaccines have limited effectiveness in controlling epidemics, aside from reducing mortality. This is evident in the UK, Israel, and the US (Fig. 6ACD), where vaccination is well underway. As *K* continues to rise, it is inevitable that new epidemics will appear in these countries. In fact, in all those countries, the number of confirmed cases decreased for a while but soon returned to the original level.

One of the characteristics of a country that has controlled the epidemic well is that it is able to suppress the *K* positives immediately (Fig. 7). Hence, the values of *K* frequently increase and decrease. In these countries, *I* can even be zero, resulting in the interruption of the line because *K* cannot be calculated. Additionally, in these countries, the negative peaks of *K* are as low as -0.2; therefore, the mean infectious time would have been 5 days or even shorter, suggesting quick discovery and isolation of infected people. For example, in Iceland (Fig. 7A), even if there are some *K-*positive days, they are quickly suppressed and do not cause epidemics. This trend has been observed in New Zealand (Fig. 7B), Australia, and China (Konishi 2021b). It takes constant effort to maintain this, and these countries continue to do so. Taiwan (Fig. 7C) experienced an outbreak of the highly contagious alpha variant (World Health Organisation 2021) in June 2021, with an *R*_0_ of 4. However, a rapid response quickly reduced *K* and maintained the number of infected people under control. Tottori is a Japanese prefecture (Fig. 7D), which had one outbreak of COVID-19 infections at the time of the Olympics, with *R*_0_ = 3.7 at the peak; however, *K* converged in a few days. The mean infectious time was particularly short (2 days), one of the shortest in the world (Fig. 4B). There seems to be a huge disparity between municipalities, even in the same country, as observed in Tokyo (Table S2). Naturally, there are fewer *K*-positive days in these countries and areas. A correlation was observed between the number of *K*-positive days and confirmed cases in the country (Fig. 7E). This is probably because these countries are able to maintain *a low τ*. In fact, there was a positive correlation between *τ* and the number of confirmed cases (Fig. 7F).

**Fig. 7.**
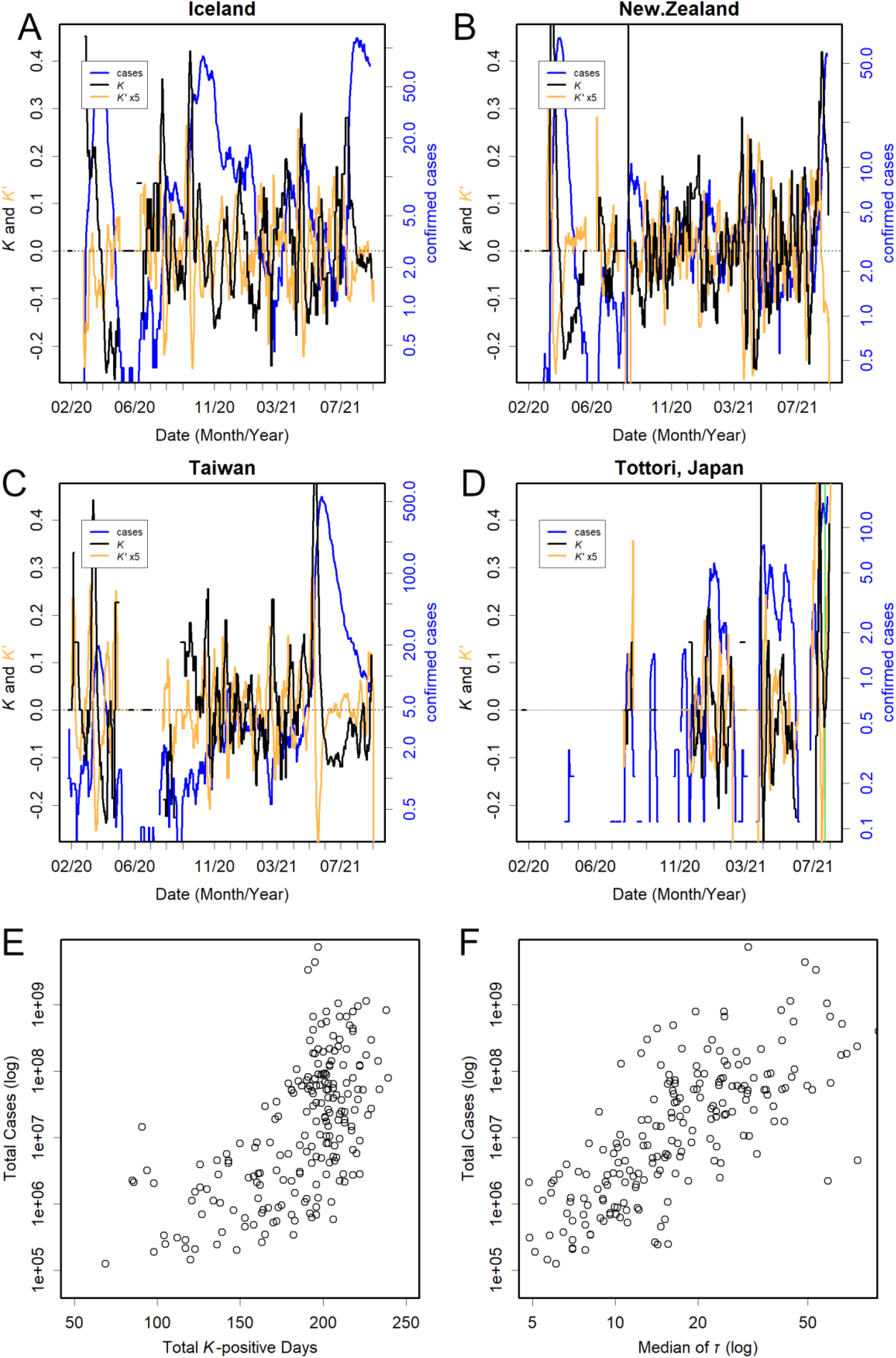
Regions with infections under control. (**A**) Iceland, (**B**), Taiwan, (**C**) New Zealand, and (**D**) Tottori, Japan. Relationships between the number of total confirmed cases and total *K*-positive days (**E**), and median of *τ* (**F**). Pearson’s corelation coefficient, *r* = 0.80 and 0.66, respectively.

In addition to *K*, mortality rate was assessed. The rate was roughly log-normally distributed (Fig. 8A). The global average was 0.017, but it is worth noting that it had an exponential spread. This value varies considerably from country to country, and mortality tends to be higher in countries where *K* does not decline (Fig. 8B and Fig. 5D). This may be due to a lack of medical care. Even in countries where vaccines are widely available, the mortality rate does not necessarily decrease as much (Fig. 8C). The rate increases or decreases with time, with the peak in mortality occurring a few months later than the peak in *I* (Fig. 8D and 8E). The number of deaths and mortality rates were lower in countries where the epidemics were well controlled, probably because they have more affluent medical resources (Fig. 8F and Table S2).

**Fig. 8.**
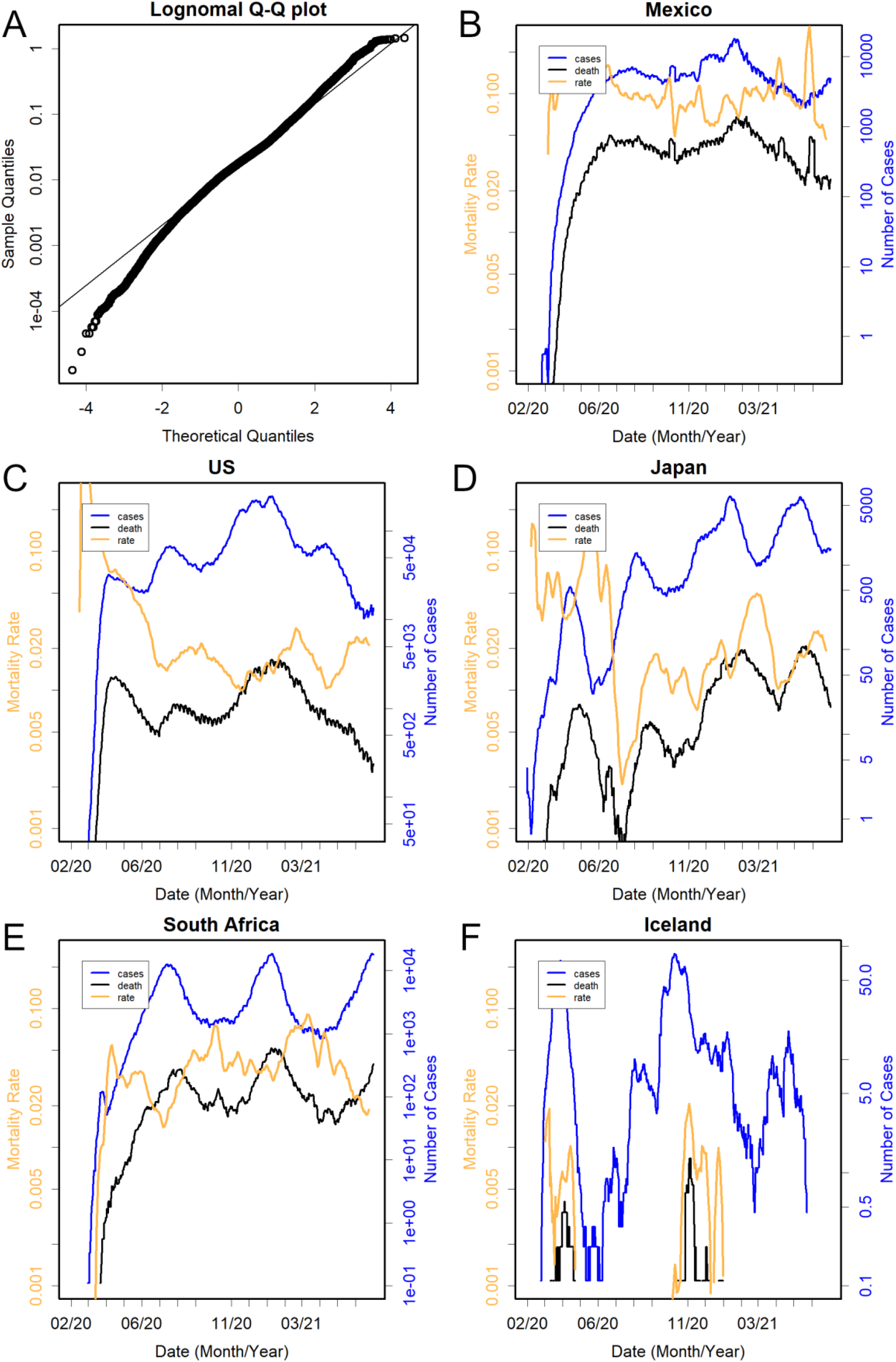
Number of deaths (black) and mortality rate. (**A**) Normal QQ plot of the logarithm of the mortality rate. (**B-F**) Situation in each country. Semi-log plot.

## Discussion

The results of the simulation, in which the infections were repeated at short intervals, showed a linear increase and decrease in *K* (Fig. 2), similar to the real data (Figs. 5–7). By observing changes in *K*, the important parameters *R*_0_ and *τ* could be estimated (Figs. 3 and 4), and by using these parameters, the SIR model could approximate changes in the confirmed cases (Fig. 4F).

These results confirm the appropriateness of this well-established model for COVID-19 epidemics. The real data suggested that the epidemics with small *S*_0_ started in a time-shifted manner with a mean interval of 11.3 days (Fig. S3). The *S*_0_ of each epidemic might be determined by the infectivity of the variant, lifestyle, and government measures.

By observing *K*, patterns of distributions of peak intervals, negative peaks of *K*, the slope of *K*, and *R*_0_ were revealed from the real data using the SIR model (Fig. 4). They were distributed according to the exponential distribution; this study was possible because *K* can be estimated simply and automatically (Table S1). Knowledge of the statistical distribution is advantageous not only for the accuracy of data analysis, but also because it provides information regarding the magnitude of the data as well as the hidden mechanism that causes the distribution (Tukey 1977). In addition, the distribution was used to determine the confidence limits of the measurements (Fig. 4).

The reason why the peak heights of *K* and *K*’ obey the exponential distribution (Fig. 4B and S2B) may be complicated. The distribution is often used to represent intervals of randomly occurring events; therefore, it appeared in the cases of the peak intervals and length of consecutive *K*-positive days (Fig. 4A and 4C). However, this distribution may also occur if variable factors increase over time, and the peak height is determined by the size of the factors when the peak occurs in a random manner (here, the timing of occurrence is unrelated to the size). A negative *K* peak was determined by how quickly a patient could be identified and isolated. Therefore, the accumulation of countermeasures may act as an increasing factor. Peaks of *K*’ correlate with the infectivity of a virus. Infectivity is determined by sequence differences; therefore, accumulated mutations may act as an increasing factor. Additionally, as *R*_0_ = 2^*Kτ*^, the logarithm of *R*_0_ would have a pseudo-exponential character (Fig. 4D). Much of the upwardly displaced data (Fig. 4D) would have been affected by the extended *τ* (Fig. 4E) rather than simply the increased infectivity (Fig. S2B). Presumably, in cases where *R*_0_ was large, medical care was more likely to be poor, and patients could not be detected and isolated in time.

We could estimate important parameters of the SIR model, such as *R*_0_ and *τ*, by observing the increase and decrease in *K* (Fig. 4). This is easier than repeating simulations to find the optimal solution and is more objective than calculating them using artificial intelligence (Rahimi et al. 2021). These approaches would estimate the parameters with higher accuracy, but this does not guarantee higher certainty. Rather, using a physical parameter, such as *K*, allows easier access to information (Table S1) and is useful for understanding and predicting epidemics, which would be beneficial for decision-making authorities.

The statistical distribution of *τ* necessitated revisions of both the isolation period and the decision to release a patient. In Japan, the isolation period is uniformly 10 days (Tokyo Metropolitan Government 2021), and no PCR test is performed when the isolation is lifted. However, the mean infectious time in Tokyo was 16 days (Fig. 5C). Furthermore, this estimated *τ* would have been shortened by the effort to find and isolate the patients; the period in which a patient excretes the virus must be longer than *τ*. Therefore, the isolation period must be longer. Furthermore, according to the SIR model, the average value represents half-life. This indicates that many patients may have been infectious upon release. Thus, the required period for isolation varies from person to person, and this cannot be determined without testing.

Observing the daily changes in *K* is important for evaluating and forecasting the state of epidemics (Figs. 7E and 7F). If *R*_0_ is low, many susceptible individuals will escape the infection (Fig. S1), but as the original *S*_0_ is unknown, information on *R*_0_ is not useful for predicting the scale of infection. If appropriate measures are taken, the epidemic converges rapidly, and vice versa, regardless of *R*_0_. Unfortunately, the magnitude of *S*_0_ did not alter the slope of *K* (Fig. 2B). In fact, while *K* and *K*’ were presented on the same scale among countries, the confirmed cases differed significantly (Fig. 5-7). Therefore, a single measurement of *K* was not useful for the evaluation. Rather, the scale of the epidemic can only be estimated from the continuous observation of the increase and decrease in *K* (Figs 5–7).

The mean value of *R*_0_ was 2.9, suggesting that in this situation, approximately 20% of the *S*_0_ would be spared from infection, and a lower *R*_0_ should be more frequent (Figs. 4D and S3A), leaving more uninfected people (Fig. S1). However, ending the pandemic by herd immunity may not be achieved. If patients are left in a city, new variants that break the previous immunity start the next epidemic. Due to the large number of infected people, this virus is now mutating at a faster rate than the N1H1 influenza virus (Konishi 2021c). Termination of the pandemic is not expected, given that the flu has not yet been terminated by herd immunity. Additionally, even small differences may find a new set of *S*_0_. In fact, approximately two-thirds of adults in India and Brazil are reported to have antibodies against the coronavirus (Deutsche Welle 2021), but the outbreak has not ended at all in these countries (Fig. 5, Konishi 2021b).

If *K* is consecutively positive for more than, for example, 10 days (Fig. 4C), the number of patients will clearly increase; if the number of *I* is already high, it is almost inevitable that there will be a peak a few weeks to a month from that time. The longer *K* is positive, the more the number of *I* increases exponentially, resulting in more cases (Fig. 7E). For example, in the case shown in Fig. 2B, *I* increased tenfold after *K* remained positive for only eight more days. Therefore, it is important to not allow a series of *K*-positives to occur. Therefore, policy measures must be implemented to reduce *K* at an early stage. The large epidemics in the US and India were not caused by high *R*_0_; they occurred because *K* was allowed to remain positive (Fig. 6). The large peaks were not caused by the speed of the infection, but because of the longer period of lack of infection control. In Tokyo, this number reached 70 days before and after the Olympics (Table S1). This is one of the worst records in the world (Fig. 4C).

The more rapid the detection and isolation of a patient, the shorter the mean infectious time *τ*. This can be attributed to the negative *K* peak (Fig. 2E). This value is probably 2 weeks or more if the patients are left untreated (Fig. 5 and 6). Figs. 4E and 7F show that *τ* should be maintained at a low value. The differences in *τ* depend on the measures taken by the government: more PCR testing, isolation of infected people, and proper lockdown and testing of all people in the area when *K* is increased. Only regions that have been able to follow this protocol can successfully control epidemics and maintain their status (Fig. 7). Many other countries are unable to maintain a negative *K* value, and thus the levels of *I* remain high (Fig. 5 and 6). This hides the initiation of new epidemics and provides a chance for new mutations to occur (Konishi 2021a).

The benefits of reducing *τ* are also evident in the comparison between Tokyo (Fig. 5C) and Tottori (Fig. 7D). Here, although the cities are different in size, infections per population and death rate differed by an order of magnitude, with the rates in Tokyo close to those in the US and UK, and the rates in Tottori were close to those in countries with infections under control. In particular, there was a double-digit difference in deaths per population between the two cities (Table S2). In August 2021, the positivity rate of PCR tests in Tokyo remained above 20%. This is because the number of tests was too small; in contrast, individuals participating in the Olympics were taking the tests daily (Tokyo Metropolitan Government 2021). On 4 August 2021 the government asked the medical institutions to keep all the patients who were not in critical condition at home, and Tokyo decided to embrace this policy (NHK 2021). In fact, a bed could not be found for an emergency patient in 100 hospitals in Tokyo (TBS 2021). Of the positive patients who requested ambulance transport between 2 and 8 August, 2021, 57% were sent back home as there was no hospital with the capacity to accept them. However, 7,000 medical personnel were mobilised for the Olympics (Yomiuri 2021). Such an irresponsible policy means the abandonment of disease control, including finding and isolating patients; therefore, *τ* in this region will rise further, consequently expanding *R*_0_ (Fig. 4E) and the number of cases (Fig. 7F). This huge difference is due to the policy of the local governments on what to treat as important.

Unfortunately, vaccines seem to have limited effectiveness as a means of ending the epidemic (Fig. 5). This is probably related to the fact that they are not given to younger children, and some people do not wish to be vaccinated. Furthermore, newer infective variants may break through acquired immunity. The simplest way to end the pandemic is to eliminate the virus, as in the cases of SARS and smallpox. Some of the countries with reduced *τ* have almost succeeded in doing this, but the newer variant that comes from the rest of the world is abolishing this attempt. COVID-19 elimination can only be accomplished through worldwide efforts. If the current situation cannot be improved, the epidemic will continue in countries where it is already prevalent.

## Conclusion

Simulations showed that the SIR model was effective for the COVID-19 epidemic. Accordingly, by continuously observing the logarithmic growth rate, *K*, we can predict the number of patients for the following few weeks up to a month. To control the epidemic, it is critically important to prevent *K* from remaining positive consecutively; rather, efforts must be undertaken to reduce *K* to end the peaks completely. It is essential to identify and isolate infected individuals to reduce the mean infectious time, which appears in the negative peaks of *K*. The mean infectious time was 12 days on average, but it could become more than twice if the patients were left untreated. Since this represents the half-life, the criteria for isolation, such as those adopted in Japan, need to be more restrictive.

## Data Availability

Any data can be downloaded and used freely.

https://doi.org/10.6084/m9.figshare.16551441.v1

---

**Fig. S1.**
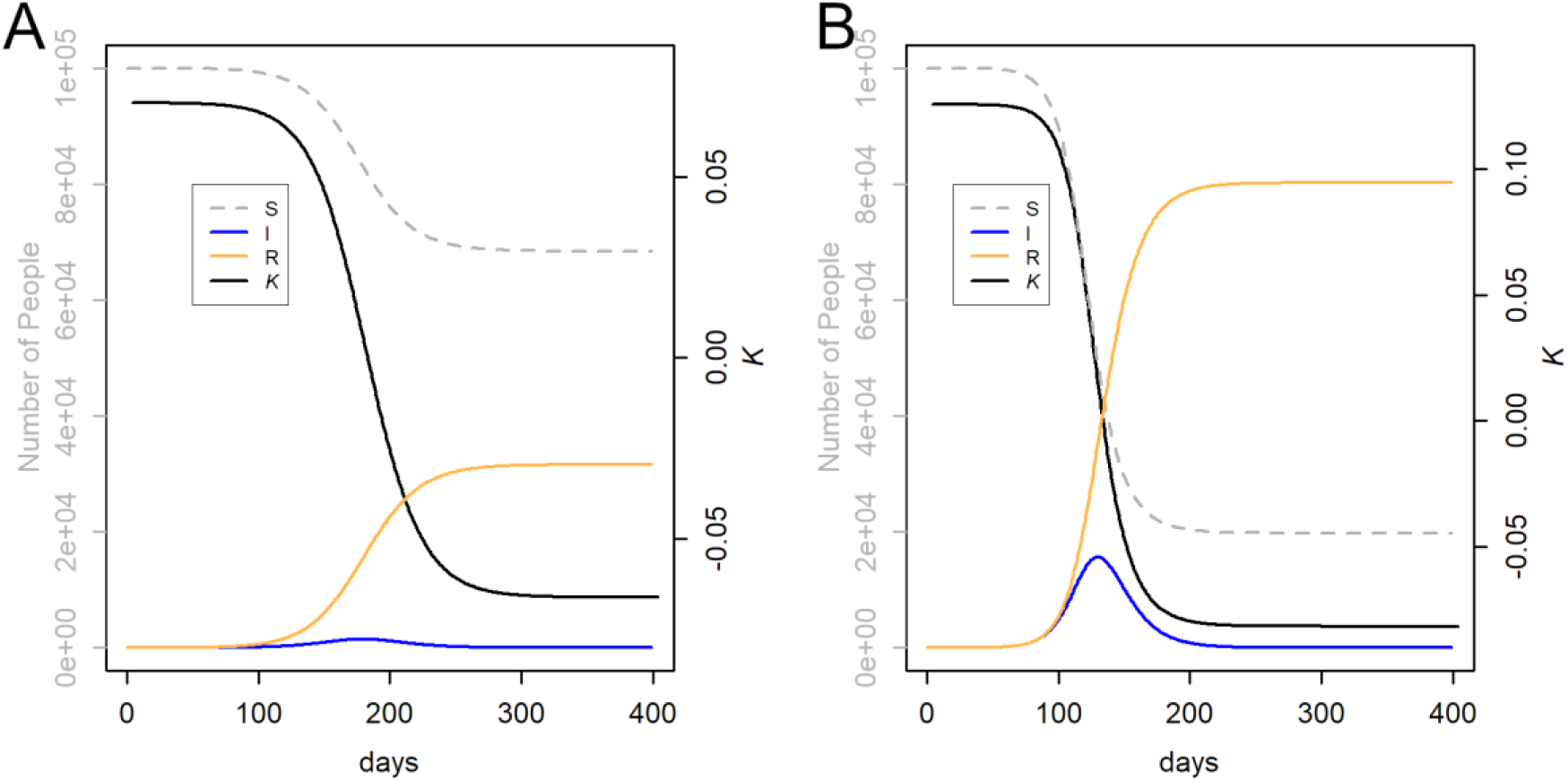
Simulation under different *R*_0_. (**A**) Close to the limit of exponential amplification: *R*_0_ = 2.1, *τ* = 15. Ca. 70% of *S*_0_ remained uninfected, and the peak of *I* remained low. (**B**) Changes under average conditions: *R*_0_ = 2.9, *τ* = 12. Ca. 20% of *S*_0_ remained uninfected.

**Fig. S2.**
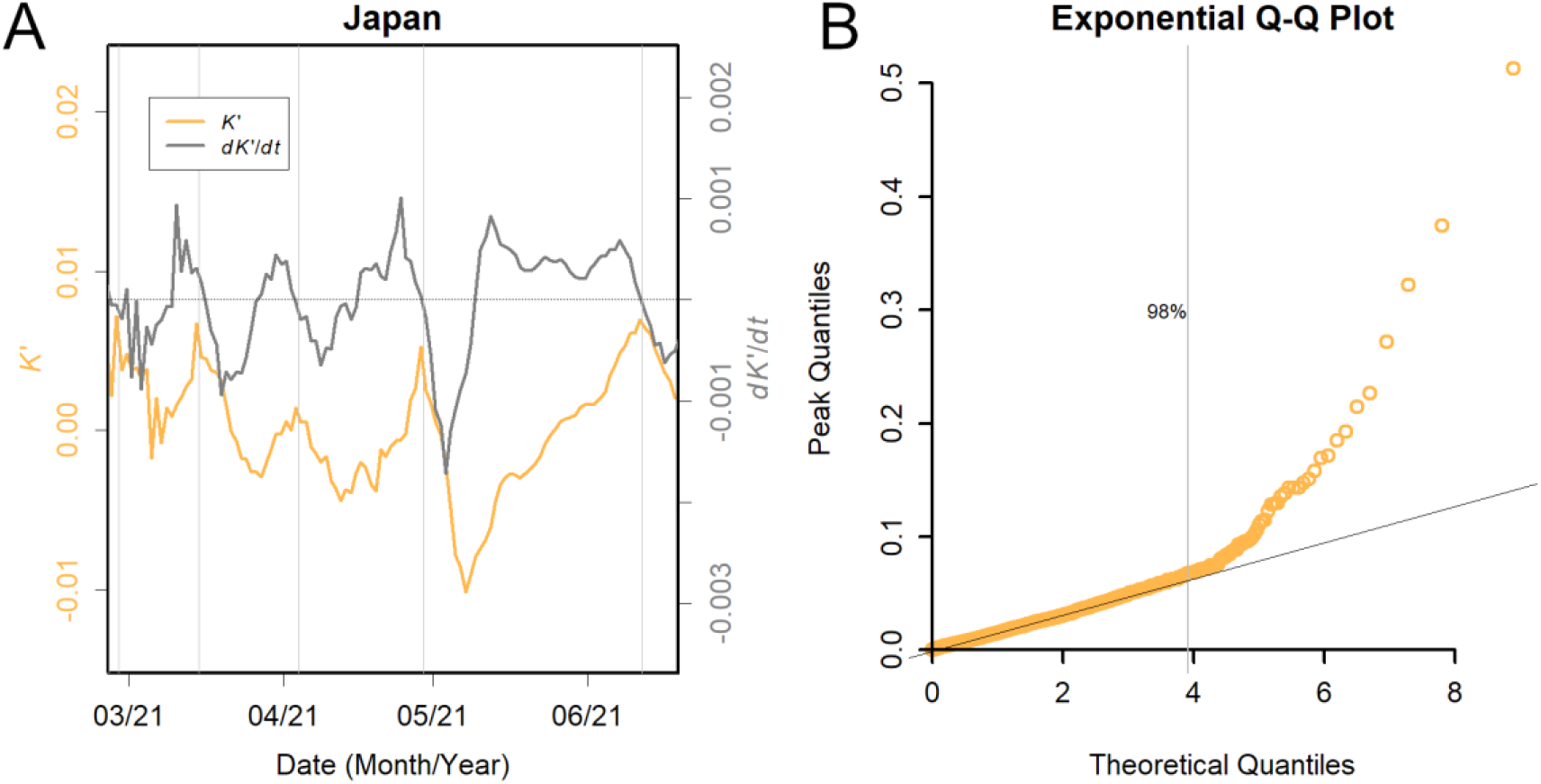
Examples of data. (**A**) Relationship between *K*’ and *dK*’/*dt*. The peak of *K*’ appears when *dK*’/*dt* becomes negative. Grey vertical lines show the positions of the found peaks. (**B**) Distribution of *K*’ peaks. Correspondence of quantiles of the peaks with that of the theoretical values. The top 2% of data may represent the effects of the super spreaders and newest infectious variants.

**Fig. S3.**
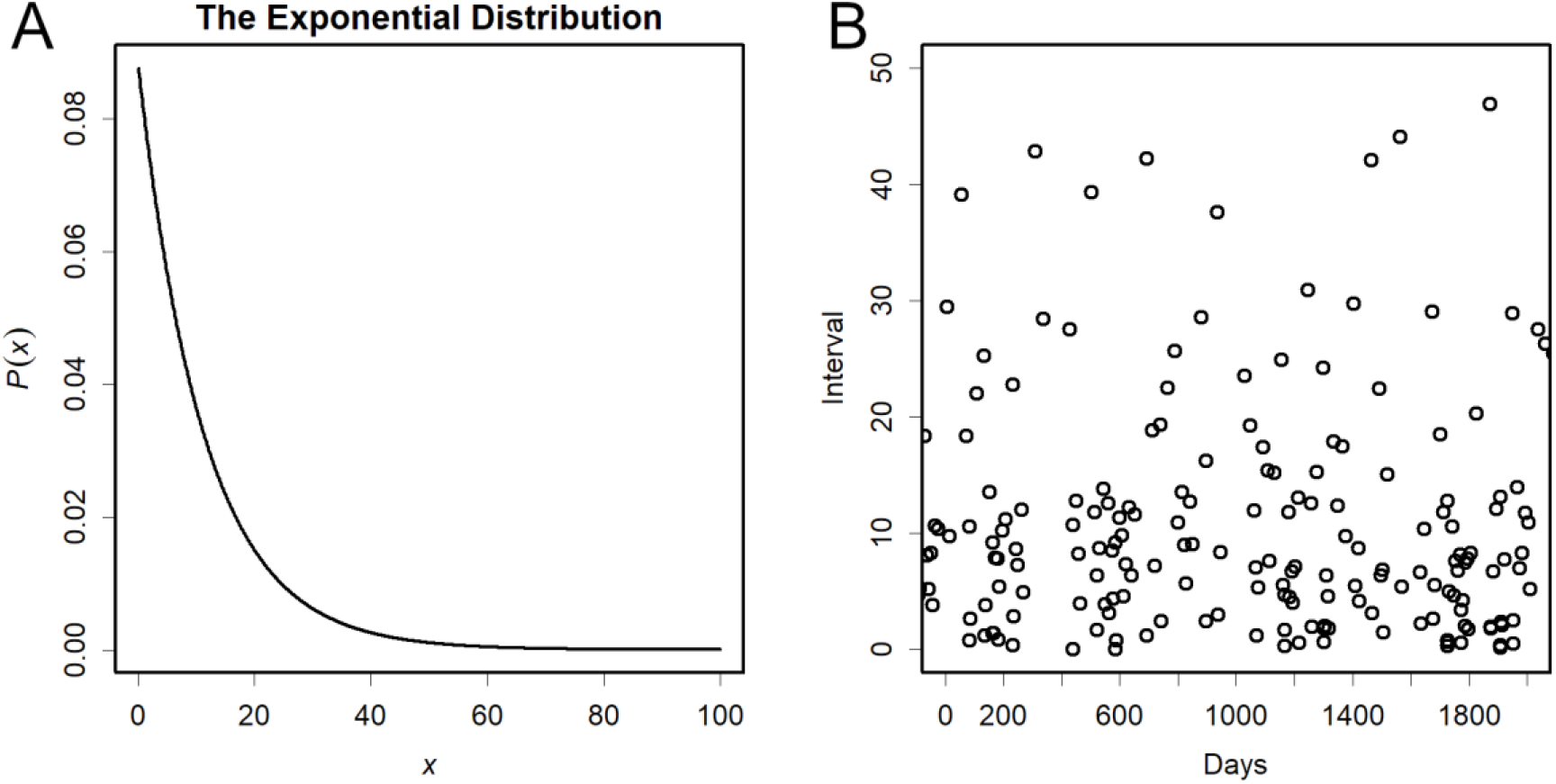
Exponential distribution. (**A**) Probability density function, *rate* = 1/11.3. The density decreases exponentially. (**B**) Frequency of intervals. A random exponential distribution with *ratio* = 1/11.3 was generated, where the vertical axis shows the respective value and the horizontal axis the total up to that value; higher values occur after such intervals of horizontal axis. Intervals of more than 40 days are observed several times over the course of 400 days.

**Fig. S4.**
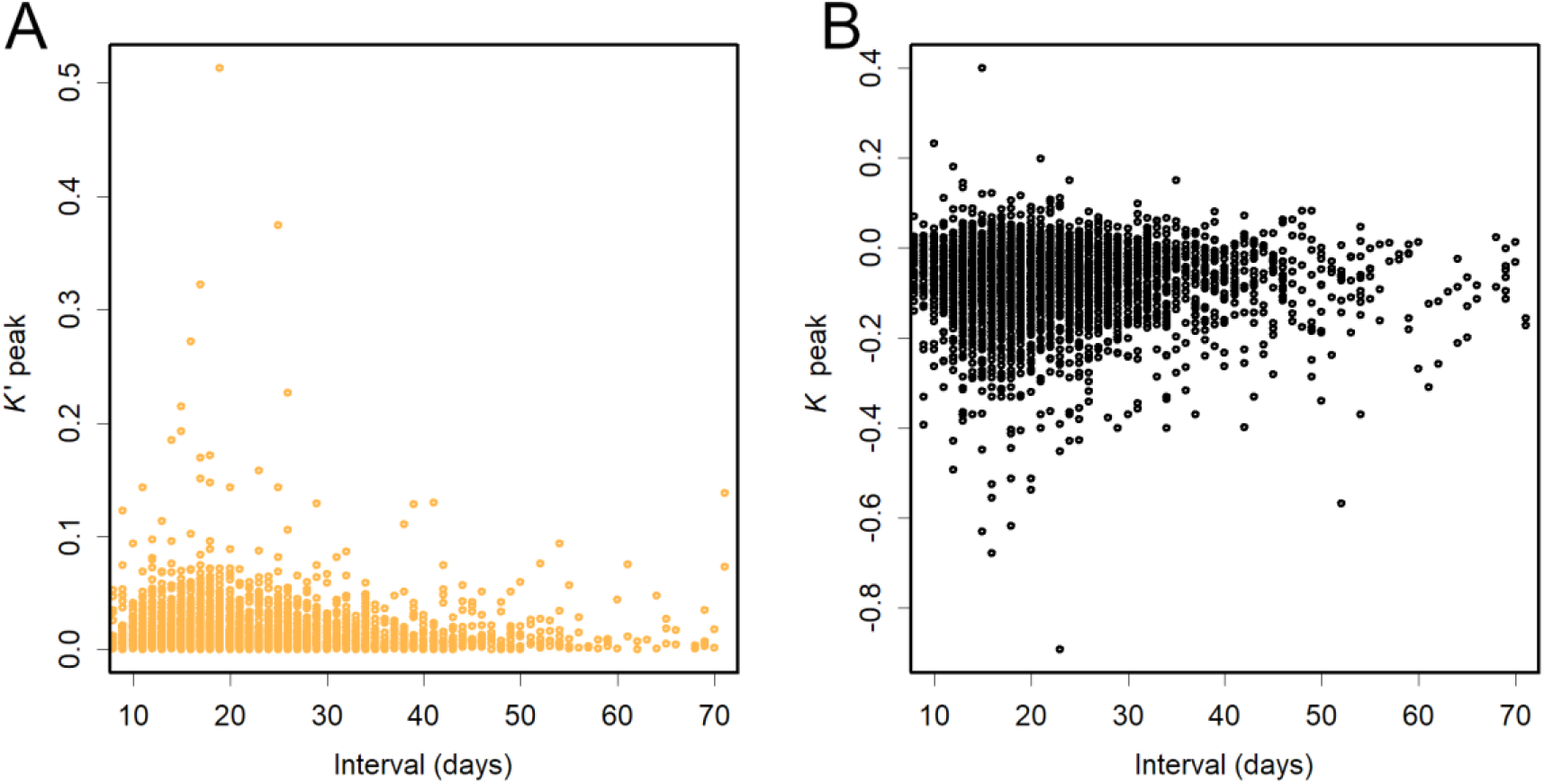
Relationship between the intervals of peaks and peak heights. (**A**) Peak of *K*’. Pearson’s correlation coefficient, *r* = -0.052. (**B**) Negative peak of *K*. Pearson’s correlation coefficient, *r* = -0.022.

**Table S1.** (TableS1.xlsx) Calculation of *K* in Tokyo. By using this table, *K* can be estimated easily.

|  | Tokyo | Tottori | Japan | England | US | Iceland | New Zealand |
| --- | --- | --- | --- | --- | --- | --- | --- |
| Mean Infectious Time | 18 | 3 | 11 | 13 | 30 | 6 | 8 |
| Population | 1.4.E+07 | 5.7.E+05 | 1.3.E+08 | 6.7.E+07 | 3.3.E+08 | 3.6.E+05 | 4.9.E+06 |
| Confirmed Cases | 1.2.E+05 | 4.9.E+02 | 8.1.E+05 | 5.0.E+06 | 3.4.E+07 | 6.6.E+03 | 2.8.E+03 |
| Death | 2.2.E+03 | 2.0.E+00 | 1.5.E+04 | 1.3.E+05 | 6.1.E+05 | 2.9.E+01 | 2.6.E+01 |
| Infection / Population | 8.5.E-03 | 8.6.E-04 | 6.4.E-03 | 7.4.E-02 | 1.0.E-01 | 1.8.E-02 | 5.6.E-04 |
| Death / Population | 1.6.E-04 | 3.5.E-06 | 1.2.E-04 | 1.9.E-03 | 1.9.E-03 | 8.1.E-05 | 5.3.E-06 |
| Death / Infection | 1.9.E-02 | 4.1.E-03 | 1.9.E-02 | 2.6.E-02 | 1.8.E-02 | 4.4.E-03 | 9.4.E-03 |

Table S2

Numbers and rates of infections and deaths up to 6 July 2021. Exponential notation. The mean infectious time is the median of the series of estimated *τ*.

## References

Alene M, Yismaw L, Assemie MA, Ketema DB, Gietaneh W, and Birhan TY. 2021. Serial interval and incubation period of COVID-19: A systematic review and meta-analysis. BMC Infectious Diseases 21:257. 10.1186/s12879-021-05950-x

Deutsche Welle. 2021. India: How reliable are herd immunity claims? https://www.dw.com/en/india-covid-sero-surveys/a-58648454

Dong E, Du H, and Gardner L. 2020. An interactive web-based dashboard to track COVID-19 in real time. The Lancet Infectious Diseases 20:533–534. https://doi.org/10.1016/S1473-3099(20)30120-1

Ellis G, and Silk J. 2014. Scientific method: Defend the integrity of physics. Nature 516:321–323. 10.1038/516321a

Guan WJ, Ni ZY, Hu Y, Liang WH, Ou CQ, He JX, Liu L, Shan H, Lei CL, Hui DSC D. B, Li LJ, Zeng G, Yuen KY, Chen RC, Tang CL, Wang T, Chen PY, Xiang J, Li SY, Wang JL, Liang ZJ, Peng YX, Wei L, Liu Y, Hu YH, Peng P, Wang JM, Liu JY, Chen Z, Li G, Zheng ZJ, Qiu SQ, Luo J, Ye CJ, Zhu SY, Zhong NS, and China Medical Treatment Expert Group for COVID-19. 2020. Clinical characteristics of coronavirus disease 2019 in China. New England Journal of Medicine 382:1708–1720. 10.1056/NEJMoa2002032

Heng K, and Althaus CL. 2020. The approximately universal shapes of epidemic curves in the Susceptible-Exposed-Infectious-Recovered (SEIR) model. Scientific Reports 10:19365. 10.1038/s41598-020-76563-8

IUPAC. 2021. lifetime, τ. in Gold book. International Union of Pure and Applied Chemistry, ed. Available at https://goldbook.iupac.org/terms/view/L03515

Konishi T. 2021a. Effect of control measures on the pattern of COVID-19 Epidemics in Japan. PeerJ https://peerj.com/manuscripts/58872/

Konishi T. 2021b. Epidemic of COVID-19 monitored through the logarithmic growth rate. Dataset. https://doi.org/10.6084/m9.figshare.16551441.v1

Konishi T. 2021c. Progressing adaptation of SARS-CoV-2 to humans. bioRxiv: The preprint server for biology:2020.2012.2018.413344. 10.1101/2020.12.18.413344

Lauer SA, Grantz KH, Bi Q, Jones FK, Zheng Q, Meredith HR, Azman AS, Reich NG, and Lessler J. 2020. The incubation period of coronavirus Disease 2019 (COVID-19) from publicly reported confirmed cases: Estimation and application. Annals of Internal Medicine 172:577–582. 10.7326/M20-0504

Li Q, Guan X, Wu P, Wang X, Zhou L, Tong Y, Ren R, Leung KSM, Lau EHY, Wong JY, Xing X, Xiang N, Wu Y, Li C, Chen Q, Li D, Liu T, Zhao J, Liu M, Tu W, Chen C, Jin L, Yang R, Wang Q, Zhou S, Wang R, Liu H, Luo Y, Liu Y, Shao G, Li H, Tao Z, Yang Y, Deng Z, Liu B, Ma Z, Zhang Y, Shi G, Lam TTY, Wu JT, Gao GF, Cowling BJ, Yang B, Leung GM, and Feng Z. 2020. Early transmission dynamics in Wuhan, China, of novel coronavirus–infected pneumonia. New England Journal of Medicine 382:1199–1207. 10.1056/NEJMoa2001316

Menon S, and Goodman J. 2021. India Covid crisis: Did election rallies help spread virus? Available at https://www.bbc.com/news/56858980.BBC.

Ministry of Health Labour and Welfare. 2021. Open data. Available at https://www.mhlw.go.jp/stf/covid-19/open-data.html

NHK. 2021. Tokyo metropolitan government considers reviewing hospitalization standards in response to government policy of “home treatment”. Available at https://www3.nhk.or.jp/news/html/20210804/k10013180591000.html

Norris K, and Gonzalez C. 2020. COVID-19, health disparities and the US election. EClinicalmedicine 28:100617. https://doi.org/10.1016/j.eclinm.2020.100617

Our world in data. 2021. Coronavirus (COVID-19) vaccinations. Available at https://ourworldindata.org/covid-vaccinations

Pandey G. 2021. India Covid: Kumbh Mela pilgrims turn into super-spreaders. Available at https://www.bbc.com/news/world-asia-india-57005563.BBC.

R Core Team. 2020. R: A language and environment for statistical computing. Available at https://www.R-project.org/. Vienna, Austria: R Foundation for Statistical Computing

Rahimi I, Chen F, and Gandomi AH. 2021. A review on COVID-19 forecasting models. Neural Computing & Applications: 1–11. 10.1007/s00521-020-05626-8

Soetaert K, Petzoldt T, and Setzer RW. 2010. Solving Differential Equations in R: Package deSolve. Journal of Statistical Software 33:25. 10.18637/jss.v033.i09

Stuart A, and Ord JK. 2010. Kendall’s advanced theory of statistics, Volume 1. Distribution theory, 6th edition. Wiley

TBS. 2021. About. Available at https://news.tbs.co.jp/newseye/tbs_newseye4327990.htm, Volume 100 hospitals in Tokyo refused to transport Corona emergency patients for 8 hours0

Tokyo Metropolitan Government. 2021. Information about the new coronavirus infection (COVID-19). Available at https://www.metro.tokyo.lg.jp/tosei/tosei/news/2019-ncov.html

Tukey JW. 1977. Exploratory data analysis. London. Reading, Mass.: Addison-Wesley Pub. Co

WHO. 2021. Tracking SARS-CoV-2 variants. Available at https://www.who.int/en/activities/tracking-SARS-CoV-2-variants/

Yomiuri Shinbun. 2021. More than half of Tokyo’s ambulance calls fail to reach patients in 959 cases.not enough hospital beds. Available at https://www.yomiuri.co.jp/national/20210819-OYT1T50245/

